# A more compact photoacoustic imaging system to detect periodontitis

**DOI:** 10.1101/2021.11.02.21265776

**Authors:** Lei Fu, Chen Ling, Zhicheng Jin, Jessica Luo, Jorge Palma-Chavez, Zhuohong Wu, Jingcheng Zhou, Jiajing Zhou, Brian Donovan, Baiyan Qi, Aditya Mishra, Tengyu He, Jesse V. Jokerst

## Abstract

Photoacoustic imaging has shown value in periodontal disease, but the large size of conventional photoacoustic transducers prevents imaging of more posterior teeth, i.e., molars. Here, we report a clinical “hockey stick”-style transducer repurposed for photoacoustic (PA) via an integrated fiber bundle. The shape of the hockey stick transducer facilitates imaging of the 1st pre-molars in contrast to conventional photoacoustic transducer designs. This tool was then deployed for photoacoustic imaging of periodontal disease and the periodontal pocket via a food-grade contrast agent (cuttlefish ink). We characterized the resolution and imaging range and then validated the system with a swine model and human subjects. We could image four additional teeth per quadrant with the smaller design versus a commercial photoacoustic transducer. Three raters evaluated the performance of the hockey stick transducer. The measurements between the probing and the PA methods were blinded, but the outcomes were highly correlated. We showed a bias of ∼0.3 mm for the imaging-based technique versus conventional probing. In addition, the inter-reliability was over 0.60 for three different raters of varying experience suggesting that this approach to evaluating dental health is teachable and reproducible. Finally, we demonstrated the utility in a human subject and can image teeth much more posterior in the mouth than with conventional photoacoustic transducers.

## 1. Introduction

Periodontitis remains a significant public health concern. According to the Office of Disease Prevention and Health Promotion, 37.4% of adults aged 45 to 74 years had moderate or severe periodontitis in 2013-2014 [1]. Periodontitis has different rates by groups: African-Americans (60.3%) versus white Americans (30.8%); less than high school degree (62.9%) versus more than a high school degree (28.8%). Periodontitis has also been linked to diabetes [2], infective endocarditis [3], cardiovascular disease [4], respiratory disease [5], and mental illness [6]. Periodontitis is a chronic inflammatory disease caused by subgingival bacteria that destroy the teeth’s supporting structures [7]. Periodontitis leads to bone loss, bone mobility, gingival inflammation, and gingival recession [8].

The extent of periodontitis is measured to indicate treatment and monitor response to therapy. Bone loss can be measured on radiographs [9], and mobility is measured via manual manipulation [10]. The degree of inflammation is subjective and is based on the color of the gingiva and the frequency of bleeding on probing [11]. Recession is the main characteristic of periodontitis and is associated with clinical attachment loss (CAL) [12]. CAL is assessed by measuring the gingival margin and the pocket depth. Dentists generally use a periodontal probe such as a Williams probe to measure pocket depth [13]. However, periodontal probing can be affected by the probing force, the insertion point, and the probing angulation [14, 15]. Periodontal probing can also cause bleeding, is painful to the patient, and is time consuming for the provider.

Dental ultrasound is increasingly used to characterize both hard and soft tissues in the oral cavity [16, 17]. Ultrasound imaging has been used for dental implant studies [18, 19], and high-resolution ultrasound imaging can help identify the gingival sulcus, gingiva margin, alveolar bone, alveolar bone crest, and the cementoenamel junction (CEJ) [20, 21]. Photoacoustic (PA) imaging monitors ultrasound created by incident light pulses [22, 23]. We previously showed that PA-US imaging can measure the periodontal pocket with a food grade contrast agent (cuttlefish ink) in swine and humans with good correlation to standard probing [24, 25]. However, the fundamental limitation of the existing technology is the relatively large size of the transducers, which prevents imaging molars and premolars. While there are miniaturized transducers for anorectal [26], endocranial [27], and laparoscopic imaging [28], these systems often use a low central frequency (5-7 MHz) and do not have the spatial resolution required when imaging the oral anatomy.

Here, we modified a so-called “hockey stick” transducer for a more compact photoacoustic system. We were motivated to use this commercially available transducer because of its relatively high (9 – 11 MHz) central frequency and angled design. While original designed for superficial applications such as intraoperative neural imaging and musculoskeletal applications, the hockey stick design is also consistent with imaging the molars [29] (up to 1st molar in human subjects). After integrating light-delivery fibers for photoacoustic excitation, we validated the system with *ex vivo* porcine jaws and could image as posterior as incisor #8 and pre-molar #12 of a healthy human subject.

## 2. Materials and Methods

### 2.1 Hardware

A commercially available hockey stick transducer (ATL CL15-7, Philips) received the PA signal with a central frequency of 9 MHz and bandwidth of 7-15 MHz. We used a 14-fiber bundle to repurpose the transducer for photoacoustic imaging (2.5-mm-diameter fibers). These fibers were coupled to a tunable OPO laser operating at 680 – 970 nm (OPOTek). A customized holder was 3D printed to mount the fiber bundles on the transducer at the desired illumination angle of 45 degree. The pulse energy from the fiber bundle is 22 mJ/pulse with 5-7 ns light pulses. A research ultrasound data acquisition system (Vantage; Verasonics, Inc., Kirkland, WA, USA) was used to collect and preprocess PA signals. The Vantage system has 256 channels with a sampling rate of 62.5 MHz. The frame rate is 20 Hz equal to the laser repetition rate. We also used a commercially available photoacoustic imaging system (Vevo LAZR; VisualSonics, Inc., Canada) with a 40-MHz transducer (LZ-550).

### 2.2 Ex vivo Validation

We used a pencil lead array to evaluate light focus. Twelve pencil leads (0.2 mm diameter) were put into a 3D-printed holder that keeps them parallel and 2 mm apart [30]. Swine jaws were obtained from a local abattoir. The pocket depths of natural swine jaws are below 2 mm, but periodontal pockets are usually deeper than 5 mm in humans [31]. Thus, we made artificially deep pockets ranging from 3 mm to 7 mm with a scalpel; 35 swine teeth were prepared including 15 artificial pockets. The melanin nanoparticles in cuttlefish ink (Nortindal, Spain) were used as a photoacoustic contrast agent to highlight the pocket as described previously [24, 25]. The absorption and PA spectra of the cuttlefish melanin particles were also measured previously and shown to be relatively flat absorbers [24, 25].

### 2.3 Pocket depth measurement

Pocket depth measurements were blinded between Williams probing method and photoacoustic method. The Williams probe has black ring markings at 1, 2, 3, 5, 7, 9, and 10 mm from the tip and was placed into the pocket along the direction of tooth root [16]. The PA images were collected along with ultrasound data for anatomy. The ultrasound image and PA image were collected at the same time and comprised the same imaging area. These two images were then synchronized in ImageJ [32]. The PA image shows the distribution of the cuttlefish ink and was used to measure pocket depth: The distance from the gingival margin on ultrasound to the farthest extent of cuttlefish ink in the pocket by PA. Three raters with different experiences are involved in this study. Rater 1 (LF) has 1 year of experience in analyzing periodontal images. Rater 2 (JL) has 1 month in analyzing the periodontal images. Rater 3 (JZ) was trained by Rater 1 for 30 minutes prior to making measurements.

### 2.4 Inter-Rater Reliability

We used the inter-rater reliability (IRR) [33] to qualify the agreement between each two of the three rater via inter-class correlation (ICC) [33]. We calculated ICC in case ICC (2,1); here, the “2” means that several judges are selected and each judge measures all the pocket depths; 1 stands for the reliability of a single rater [34].

### 2.5 Human Subjects Imaging

We recruited a healthy adult male in his twenties with good oral hygiene. All work with human subjects was approved by the UCSD IRB and conducted according to the ethical standards set forth by the IRB and the Helsinki Declaration of 1975. The participant gave written informed consent and incisor #8, pre-molar #12 were imaged. The volunteer and operator both wore near-infrared protective laser goggles during experiments. Cuttlefish ink contrast agent for human subject were prepared independently [24]. We used drinkable water instead of PBS as solution. An ultrasound gel pad (Parker laboratories, Inc., USA) was adapted to create a gap between the tissue and the transducer to couple light and ultrasound. Sleeves were used to cover the gel pad and transducer in human studies.

## 3. Results and Discussion

We first compared the performance features of the various transducers and describe how we converted the hockey stick transducer to photoacoustic mode. We then describe the performance metrics of that transducer, strategies to couple optical excitation, and then validate it ex vivo and in vivo.

### 3.1 Hockey stick transducer

Figs. 1(a) and 1(b) compare the three different transducers available in our lab for periodontal imaging including the 9 MHz hockey stick transducer CL15-7, 20 MHz transducer L22-14vX [35], and 40 MHz transducer LZ-550 [24, 25]. The three transducers have different shapes. The hockey stick transducer has overall the smallest width as defined in Fig. 1(a), and much lower costs than the other two transducers. The other two transducers have higher frequency and therefore higher resolution than CL15-7. Our goal was to image teeth in the posterior of the mouth, and the stars in the teeth column mark the teeth a transducer can access for imaging. The gray stars represent teeth that are accessible for ultrasound imaging, and the red star indicates teeth accessible for PA-ultrasound imaging. The hockey stick transducer can image the upper incisors, cuspids, and the 1st premolar; the LZ-550 can only image the upper incisors in PA mode.

**Fig. 1.**
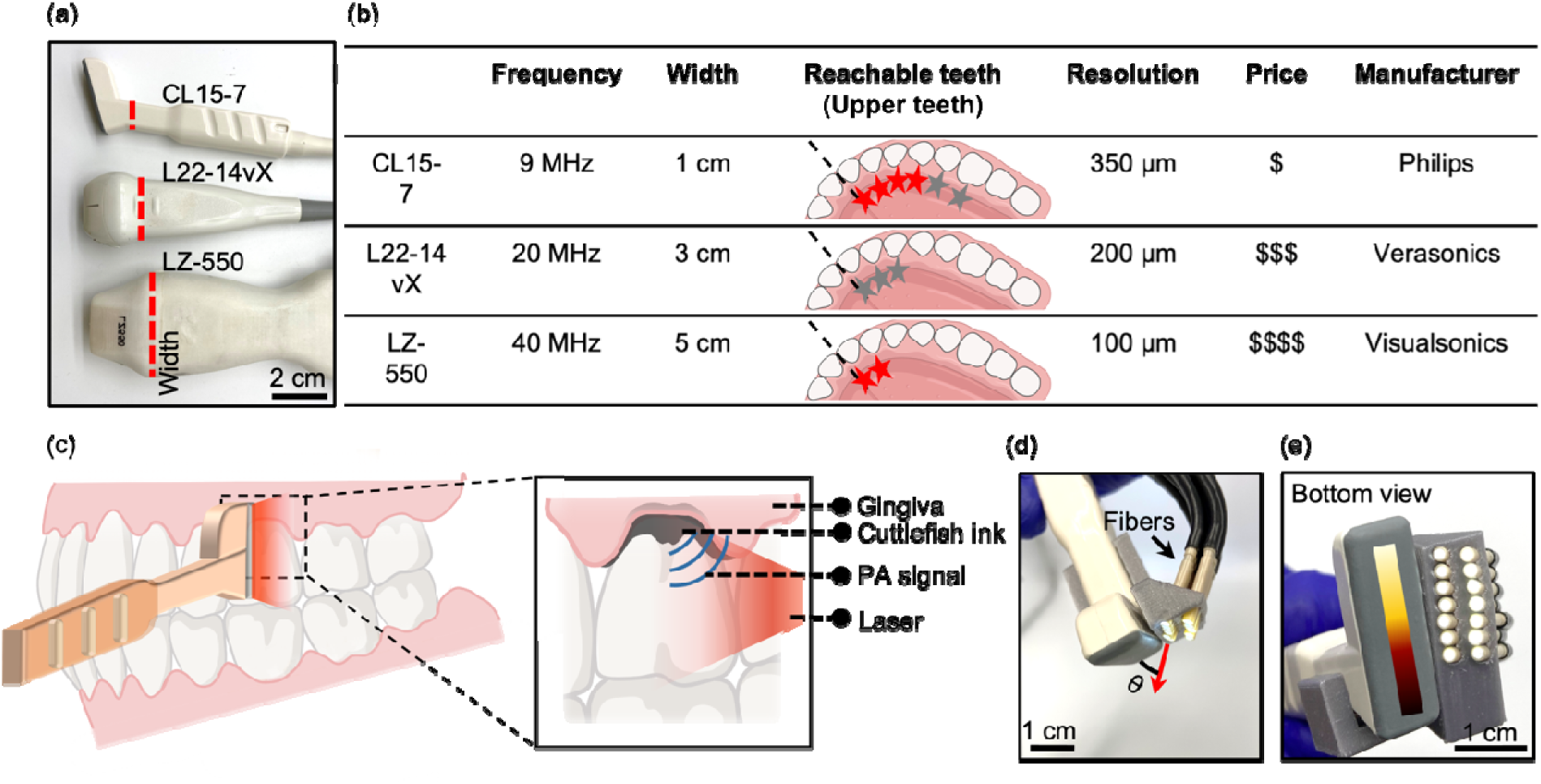
Transducer Evaluation. The smaller transducer facilitates access to four additional teeth per quadrant versus our existing photoacoustic transducer. a) A photo of three transducers, CL 15-7, L22-14vX and LZ-550. b) Performance metrics of the three transducers in (a). The stars mark the imageable upper teeth by different transducers. Gray stars stand for ultrasound imaging with red stars stand for PA-US imaging. The dashed line is the midline of human teeth. c) Photoacoustic imaging detects the periodontal pocket with a hockey stick transducer. The inset shows that cuttlefish ink to highlight the pocket. The contrast agent labels the periodontal pocket underneath the gingiva for PA imaging. d) and e) Hockey stick transducer with fiber bundle from front view and bottom view. One side of the transducer is arranged with two rows fibers while the other side has no fibers because it needs to fit into the oral cavity.

This hockey stick design was targeted for superficial high-resolution imaging including in peripheral nerves [36]. Most other clinical transducers have a lower central frequency (1 to 5 MHz) to achieve deeper penetration. The 9 MHz hockey stick transducer used here has a focal point of around 1 cm, which is shallower than other clinical transducers, but it ideally suited for periodontal applications. Fig. 1 shows the hockey stick transducer illustrating how it can be placed beneath the gums for molar imaging. The transducer surface faces the sagittal tooth surface and gingiva; the handle then projects out toward the operators. We used the CL 15-7 to obtain ultrasound images of the 1st molars (#3, #14, #19, #30) in a human subject. The periodontal pocket is usually around 5 mm below the gingival margin, which fits the near field metric of hockey stick transducer.

### 3.2 Integration of Optical Components for Photoacoustic Imaging

Figs. 1(d) and 1(e) show the front view and bottom view of the hockey stick transducer with the fiber bundle. We used 14 fibers in two rows to deliver pulse laser to the top of the transducer. Our imaging target is the ∼ 1 cm surrounding the gingiva margin, and thus we only needed to use half of the 64 elements in the 2-cm transducer. The angle of fiber orientation was 45 degrees and was chosen based on the acoustic focal zone (5 - 10 mm from the transducer) and the offset required for the coupling pad. Unfortunately, integration of the fibers added several cubic centimeters of bulk to the transducer. Thus, we only placed them on one side to reduce this bulk (Fig. 1(d)).

### 3.3 Characterization

We characterized the resolution and the imaging range of the system. The transverse and axial resolution were determined by imaging the cross-section of a tungsten wire with diameter of 25 µm (850-nm excitation via OPO laser). The PA image of the wire cross-section is shown in Fig 2(b). The transverse and axial amplitude distributions across the tungsten wire were extracted from Fig. 2(b), and the axial data are shown in Fig. 2(c). We define the FWHM (full width at half maximum) of the transverse and axial amplitude distributions as the axial and transverse resolution, respectively [37]. FWHM can be observed by doing Gaussian fitting of the amplitude distribution. The axial and transverse resolution are 320 µm and 370 µm separately, which are close to previous measurements [38, 39].

**Fig. 2.**
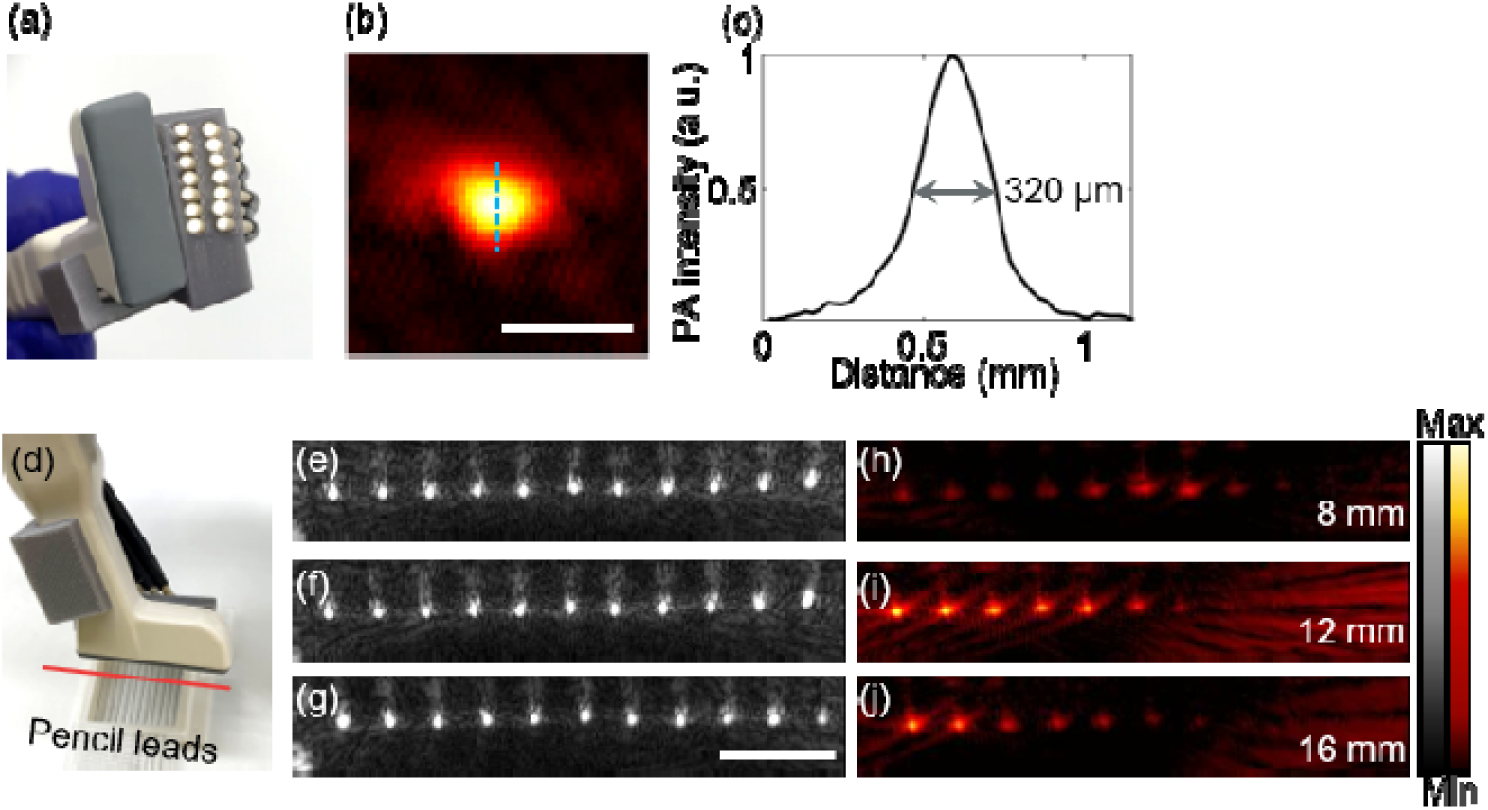
Performance Metrics of Customized Transducer. a) Overview of the hockey stick transducer and fiber array; 14 fibers deliver most light to the top of the transducer. (b) PA image of tungsten wire. (c) Axial PA amplitude distribution along the tungsten wire. 320 µm is the full width at half maximum of the amplitude distribution. It is defined as the axial resolution. (d) Schematic of pencil lead array imaging. Red line indicates imaging plane shown in e-j. Panels (e), (f), and (g) are the ultrasound images when the pencil leads array is 8 mm, 12 mm, and 16 mm below the transducer, respectively. Panels (h), (i), and (j) are the PA images when the pencil leads array is 8 mm, 12 mm, and 16 mm below the transducer, respectively. The scale bar is 5 mm.

We also imaged an array of pencil leads (12 leads, 24 mm wide) to examine the light focus as shown in Fig. 2(c). Stronger PA signal indicates better light focus. The array was placed 8 mm, 12 mm, and 16 mm underneath the transducer as shown in the ultrasound images in Fig. 2(e), 2(f), and 2(g), correspondingly. Fig. 2(h), 2(i), and 2(j) are the PA images. Obviously, the left half of the array has stronger PA signal than the right half because that is where the optical fibers are focused. The strongest PA signal is seen 12 mm beneath the transducer, which also agrees well with our settings in light focus.

### 3.4 Imaging the Williams probe in the periodontal pocket

Our previous work showed correlation between PA-based periodontal probing and Williams probing. Here, we further demonstrate that PA is a more accurate method than the Williams probing method. The Williams probe reports depths in integer values to the nearest millimeter Fig. 3(a) and 3(b). In contrast, image-based measurements have a resolution as high as 300 µm when using the hockey stick transducer and 100 µm when using a 40 MHz transducer. However, it is not clear if imaging signal/contrast agent is attenuated as a function of penetration depth. It is also not clear if PA measurement and Williams probing measurement will be performed at the same position for comparison. To answer that question, we imaged a Williams probe inside the periodontal pocket (Fig. 3(b). In other words, the Williams probe measures the pocket depth and works as ‘contrast agent’ for PA at the same time and position similar to the PA imaging of a needle [40].

**Fig. 3.**
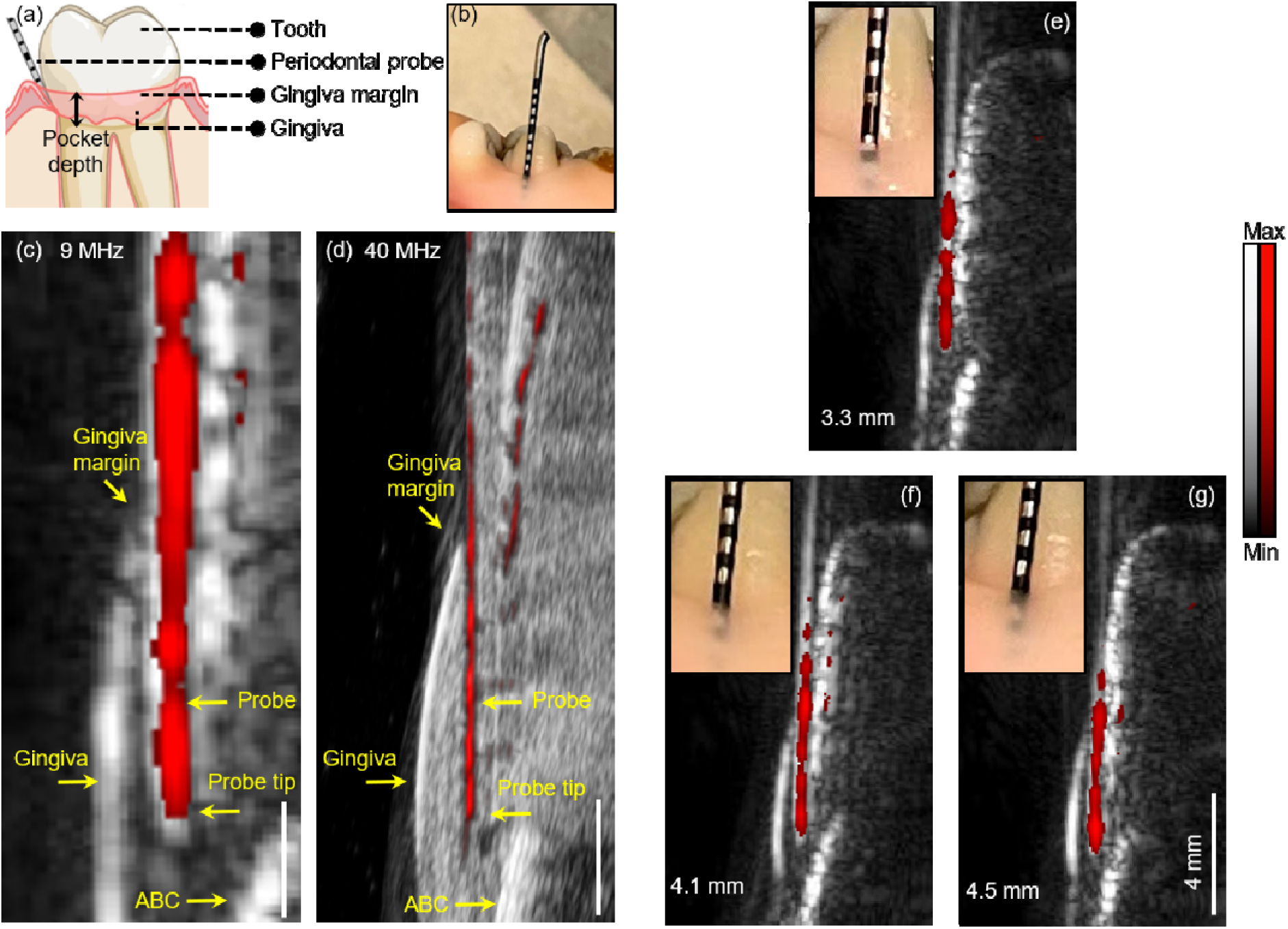
A Williams probe measures the pocket depth and offers PA contrast. a) and (b) Williams probe in the periodontal pocket with 1-mm black tick marks on the probe. c) and (d) Ultrasound/PA overlaid images of swine tooth and the Williams probe in the pocket. Panel c) is acquired with the 9-MHz hockey stick transducer. d) 40 MHz LZ-550 transducer. Scale bars in c and d are 2 mm. Ultrasound image is in gray and PA image is in red. The red line is the probe. The alveolar bone crest (ABC), gingiva, gingiva margin, and probe tip in the pocket are obvious. Artefacts under the probe in panel (d) are due to the probe and their dramatically different speed of sound. Panels (e), (f), and (g) are a stack of PA-US images when the probe is inserted into the pocket at probing depths of 3.3 mm, 4.1 mm, and 4.5 mm, respectively, with the hockey stick transducer. The insets are zoomed photos of the probe in the pocket.

Fig. 3(c) and 3(d) show the PA-US images acquired by the 9 MHz hockey stick transducer and the 40 MHz LZ-550 transducer, respectively. As expected, the 40-MHz transducer provides much better resolution than the 9-MH transducer. Nevertheless, the structure of tooth is still distinguishable with the 9-MHz transducer Fig. 3(c). We ca identify the Williams probe as a long red line in the PA image. Part of the Williams probe is in the pocket. Fig. 3(c) and 3(d) also show the gingival margin and the probe tip. We measured the distance between the gingiva margin and the tip as the probe length in the pocket. Fig. 3(e) to (g) are PA images when the Williams probe was inserted int the pocket from around 3.0 mm to 4 mm. The insets show the probe in the pocket when the PA-US image is acquired. We qualified the three probe lengths in Fig. 1(e-g) with 5 replicates: 3.3 mm, 4.1 mm, and 4.5 mm separately. In comparison, the probing depths are 3 mm, 4 mm, and 4 mm by eye, which is less precise than PA method as the probe was being inserted deep into the pocket.

### 3.5 Periodontal Pocket Depth Measurements

#### 3.5.1 Pocket Depth Measurements in swine model

We used swine tooth model to validate the performance of the hockey stick transducer because swine tooth have similar structures as human tooths [41]. We first used a Williams probe to measure the pocket depth and then performed photoacoustic imaging to measure the same pocket for comparison. Ultrasound/PA images were also acquired before applying the cuttlefish ink contrast agent as control images.

Fig. 4(a) shows the ultrasound/PA images of two swine teeth: the 4th pre-molar and the 2nd molar. PA image in red is overlaid with the ultrasound image in gray. As shown in the top two images in Fig. 4(a), the structure of swin tooth and its associated tissue are distinguishable in the ultrasound images such as gingiva, gingiva margin, occlusal surface, and ABC. As shown in the bottom ultrasound/PA images, the contrast agent is shown as a line below the gingiva margin, i.e., the periodontal pocket. The periodontal pocket is deeper in the 2nd molar than 4th pre-molar. In comparison, the top images have no PA signal in the pockets when cuttlefish ink contrast agent is not used. This confirms that the cuttlefish ink can be a contrast agent to image the pocket. By drawing a line between the gingiv margin in the ultrasound image and the deep end of the contrast agent in the PA image, we measured the pocket depth to be 1.2 mm for the 4^th^ pre-molar and 5 mm for the 2^nd^ molar.

**Fig. 4.**
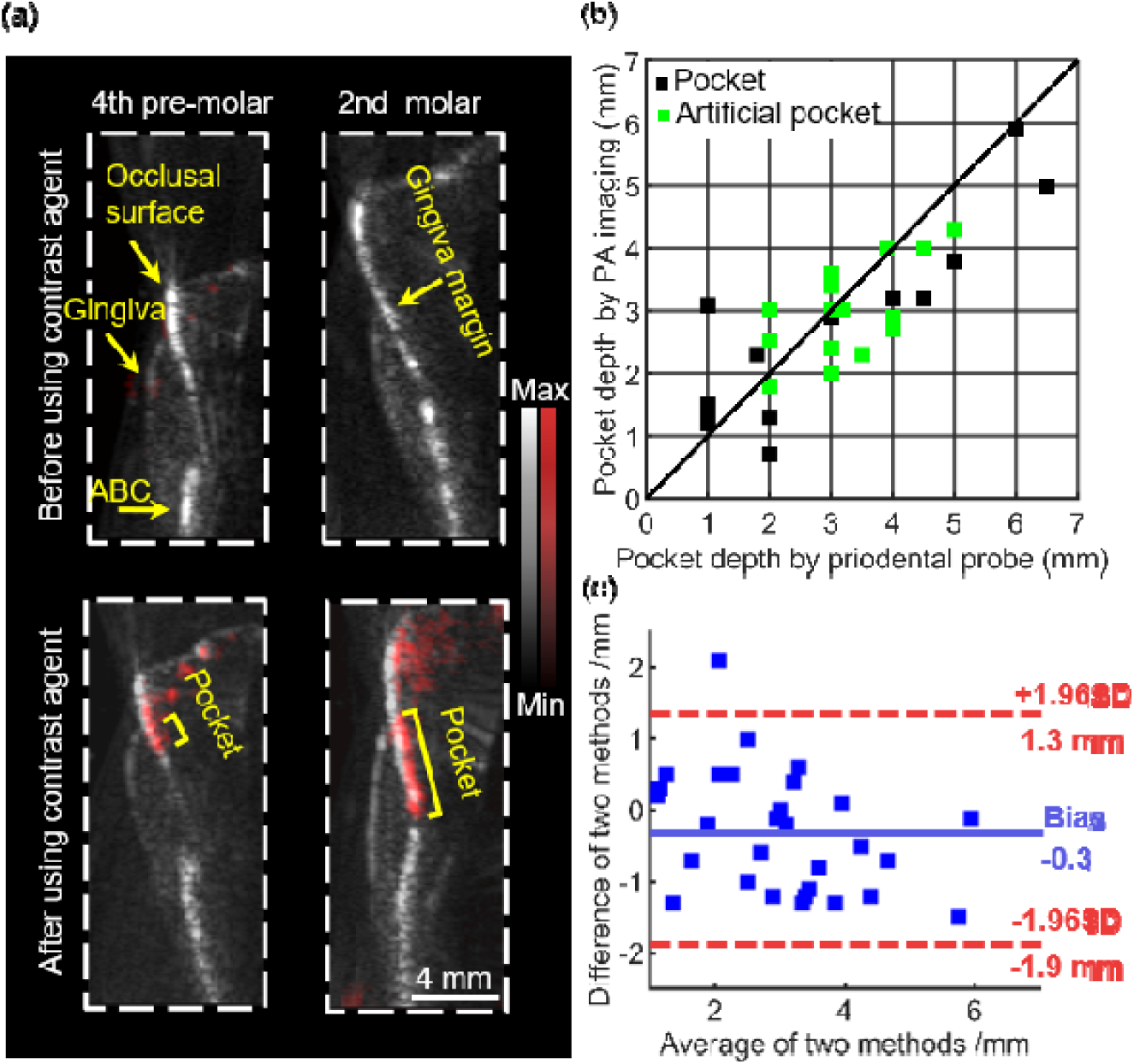
a) Ultrasound/PA images of two swine teeth, the 4th pre-molar and the 2nd molar in sagittal view. PA data are in red. Ultrasound data are in gray. Data on top are before administering the cuttlefish ink, and data on the bottom are after; hence, there is red signal. (b) Pocket depth measurements with 35 replicates. The X-axis represents the measurements with Williams probing, and the Y-axis represents the measurements with PA imaging. Black data points are the natural pockets and green data points are the artificial pockets. (c) Bland-Altman analysis of the statistics in (b). Blue line is the bias of PA measurements over probing measurements, which is -0.3 mm. The two red lines are the upper and lower limits of agreements (LOA); 95% of the replicates fell into the region between the two lines.

We then imaged 35 swine teeth for replicates and had three different raters to measure the pocket dept independently. The X-axis represents the Williams probing method, and the Y-axis represents the PA method. Blac data points are the natural pockets and green data points represent the artificial pockets (Methods). The differences of the two measurements are much less than 1 mm, and most data are close to the convergent line. Fig. 4(c) is the Bland-Altman analysis of Fig. 4(b), which qualifies the agreement between the PA and the Williams probin methods. It shows that 95% of the replicates fell within 3.2 mm of the differences between the two methods [42]; the bias is -0.3 mm. The bias can be understood as the pocket depth goes deeper, the contrast agent cannot penetrate through as these PA measurements are less than the probing measurements as shown in Fig. 4(b) [25, 43].

Table 1 shows the Bland-Altman analysis of the three raters. Rater 2 has the smallest bias of -0.06 mm while Rater 1 has the smallest LOA. The negative bias was consistent across the three raters. This is likely because PA measurements become less accurate at a greater depth due to greater interference from increasing gingival thickness. We quantified the agreement level between each two of the three raters by IRR, which is estimated by calculating the ICC (Methods). As a rule of thumb, Cicchetti (1994) provides commonly-cited cutoffs for qualitative ratings of agreement based on ICC values with IRR being poor for ICC values less than 0.40, fair for values between 0.40 and 0.59, good for values between 0.60 and 0.74, and excellent for values between 0.75 and 1.0 [33].

**Table 1.**
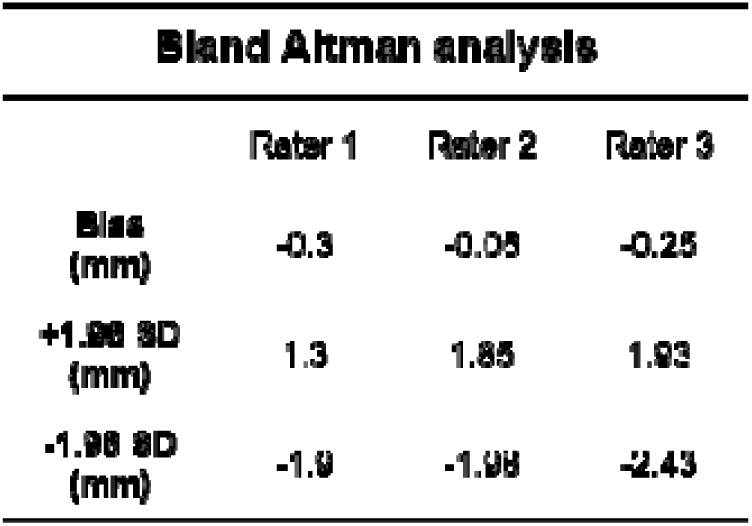
Bland-Altman analysis of the three raters. The three raters measured the 35 pocket depths independently (by imaging). The values from Williams probe were constant and were measured by an investigator blinded to the imaging results. Rater 1 has 1-year experience in PA periodontal imaging. Rater 2 has 1-month experience in PA periodontal imaging. Rater 2 was trained by rater 1 for 30 mins before doing measurements. The negative bias was consistent across the three raters. This is likely because PA measurements become less accurate at greater depths due to greater interference from increasing gingival thickness.

Table 2 shows the intra-class correlations (ICC) between Rater 1 and Rater 2, Rater 1 and Rater 3, Rater 2 and Rater 3. The agreement between Rater 1 and 2 is 0.9 (excellent). The agreements between Rater 1 and 3 as well as Rater 2 and 3 are 0.64 (good) and 0.61 (good), respectively. The differences of agreement make sense because rater 3 had been trained by rater 1 for only 30 min. Similar to clinical ultrasound imaging, interpreting the PA/ultrasound images of tooth and the associated structure requires specific expertise and is a major challenge. Previously, the ICC for dental students was 0.51 (95% CI: 0.33 to 0.75) and 0.41 (95% CI: 0.24 to 0.64) for dental faculty [44]. Our value reaches a clinical significance level is shown to be at least at a “good” suggesting that our newly developed method is systematic and teachable. The definition of excellent and good are given in Methods.

**Table 2.**
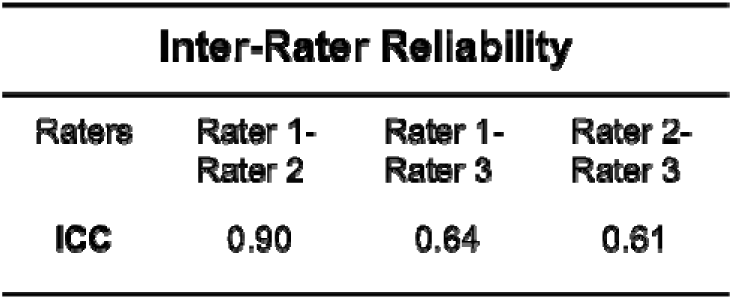
Inter-rater reliability between each two of the three raters. ICC stands for inter-class correlation (ICC). ICC is one way to qualify the inter-rater reliability (Methods). Rater 3 has the least experience in analyzing the PA/ultrasound images. As a rule of thumb, Cicchetti (1994) provides commonly-cited cutoffs for qualitative ratings of agreement based on ICC values, with IRR being poor for ICC values less than 0.40, fair for values between 0.40 and 0.59, good for values between 0.60 and 0.74, and excellent for values between 0.75 and 1.0 [33].

#### 3.5.2 Pocket Depth Measurements in a human subject

We also used hockey stick transducer to image the periodontal pocket of human teeth. The frame rate is 20 Hz, which is higher than our previous work (6 Hz) [24] and can help solve issues related to subject motion and probe stabilization. We also used water instead of ultrasound gel to couple the transducer to the teeth and gums, which improves clinical experience. However, the SNR and resolution of our system is lower than the 40 MHz system.

We imaged two teeth of a healthy human subject: incisor #8 and pre-molar #12 as shown in Fig. 5. The contrast agent was first administrated to the teeth (left panels) and then rinsed off (right panels) as control. All the images are in sagittal view. The ultrasound images show human teeth have similar structures with swine teeth. Gingiva, occlusal surface and the pocket are distinguishable in the ultrasound images. PA images in the left panels show that the contrast agent labels the pocket as well as the gingiva and occlusal surface, where the periodontal pockets and the gingiva margins can be identified in the PA images. We measured the pocket depths as 0.9 mm and 0.5 mm, which are close to the results of human subject in our previous work [24]. Healthy subjects generally have lower pocket depths than periodontal cases. After the contrast agent was rinsed off (right panels), US images still shows the structure of teeth while the PA signal is dramatically decreased, which proves that the contrast agent did stain the pocket, gingiva surface, or the occlusal surface.

**Fig. 5.**
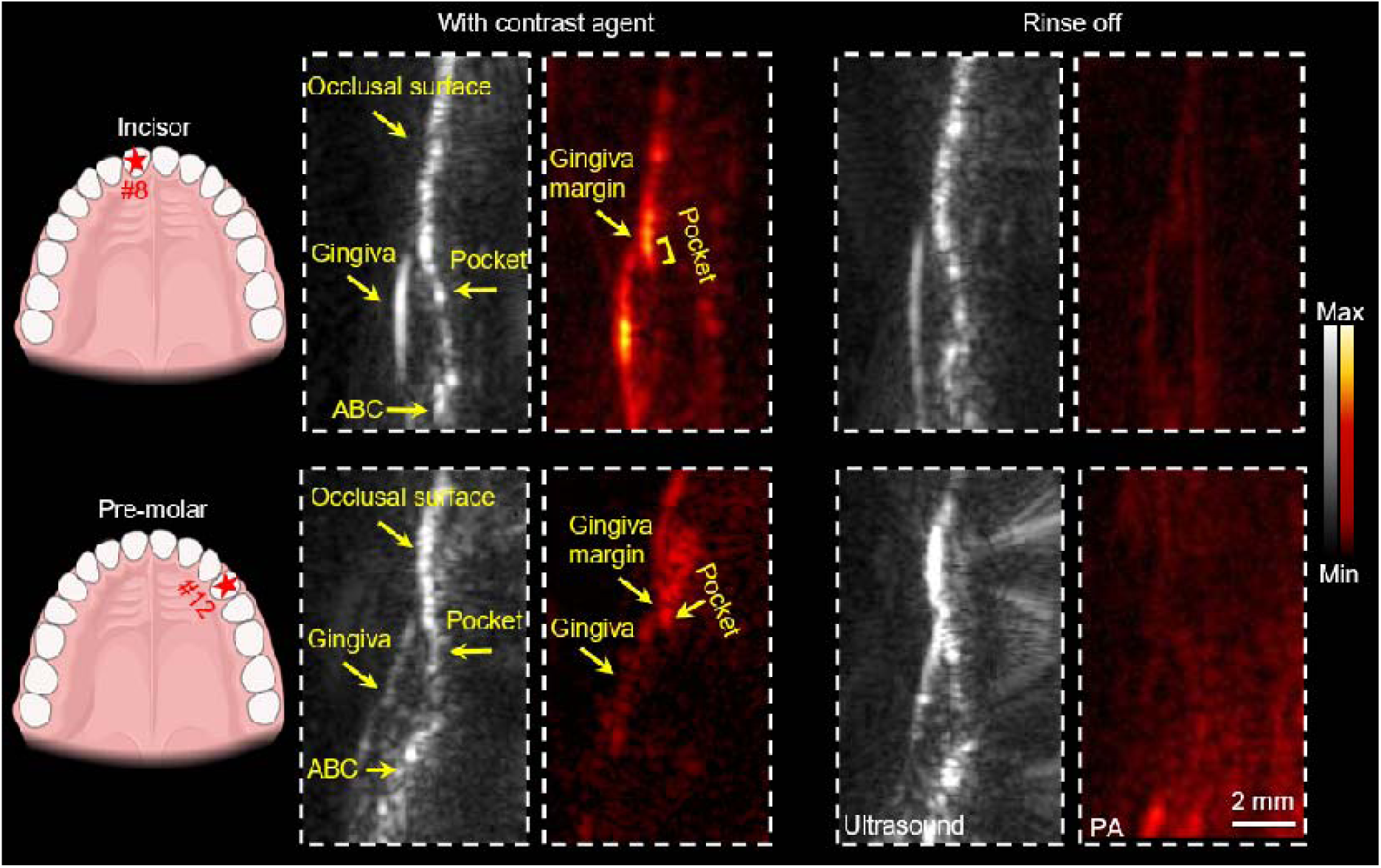
Ultrasound/PA images of two human teeth, incisor #8 (top), and pre-molar #12 (bottom). Cuttlefish ink contrast agent labels the periodontal pockets (left panels). The contrast agent is rinsed off (right panels). Ultrasound images are in gray and PA images are in red. Ultrasound images show the common structure of tooth, PA images show the position of contrast agent.

This hockey stick transducer still has some limitations. First, the last two molars of human subjects are not imageable in PA mode due to the large size of the device. The illumination angle from the fibers requires around 1.0 cm gap between the transducer and the tissue for light coupling. This gap and the size of the fiber module prevent the transducer from covering all the teeth. A new illumination method is needed. Second, high-frequency transducers with smaller dimensions that better fit the human oral cavity are needed. The periodontal imaging depth (gingival thickness) is <5 mm, which allows use of a high-frequency transducer. The periodontal pocket can be as deep as 10 mm. Thus, an even smaller linear array could be used and still cover all teeth in clinical studies.

## 4. Conclusion

In conclusion, the unique angle shape of hockey stick transducer allows it to cover more teeth than regular linear transducers. This study mainly demonstrated the ability of hockey stick transducer to detect the periodontal pocket in photoacoustic imaging. The measurements by using PA and Williams probe agree well with each other in swin model. We used hockey stick transducer to image incisor #8 and pre-molar #12 of a healthy human subject. We envision a more compact high-frequency transducer that is designed specifically for dental imaging to cover all the teeth of any human subject.

## Data Availability

All data produced in the present study are available upon reasonable request to the authors

## 5. Back matter

### 5.1 Funding

This study was funded by National Institutes of Health under grants R21 DE029025, R21 DE029917, and R21 AG065776. We also acknowledge infrastructure under grant S10 OD021821.

### 5.2 Disclosures

J.V.J. is a co-founder of StyloSonics, LLC.

### 5.3 Data availability

Data can be requested from the authors at any time.

